# A rare-variant analysis of *FADS*-related metabolites in Major Depression

**DOI:** 10.64898/2026.09.29.26364222

**Authors:** Laurence Nisbet, Ella Davyson, Robin Beaumont, Jess Tyrrell, Xueyi Shen, Andrew M McIntosh

**Affiliations:** Institute of Neuroscience and Cardiovascular Research, University of Edinburgh; Institute of Genetics and Cancer, University of Edinburgh, Edinburgh, UK; Department Clinical and Biomedical Sciences, University of Exeter

**Keywords:** Major Depression, Psychiatry, Metabolites, Genetics, Rare variants, Mendelian Randomisation

## Abstract

**Background:** Genetic and observational studies have identified metabolic factors potentially contributing to the risk of Major Depression (MD). However, studies to date have concentrated on common genetic variants, many of which are in linkage disequilibrium. This makes it difficult to localise variants to a single gene. To overcome this, we used short read whole-genome sequenced datasets (srWGS) to conduct a targeted rare variant association analysis of the *FADS* region, which has previously been implicated in metabolic effects on MD.

**Methods:** Using srWGS data from 740,373 individuals from All of Us (AoU) and the UK Biobank (UKB) we tested for associations between rare variants in the *FADS* region and MD. Additionally we conducted association tests between rare *FADS* variants and five blood-based metabolites implicated by previous studies of MD. Finally, we applied Mendelian randomisation to identify causal effects of metabolites on MD using rare-variants.

**Results:** We found *FADS* rare variants were significantly associated with all tested metabolites, but not with MD. Using Wald-Ratio MR we found a putative causal effect of Docosahexaenoic acid (DHA) on MD using a rare-variant instrument in the *FADS2* gene.

**Conclusion:** Our study represents the largest rare-variant analysis of MD to date. However, the lack of associations between *FADS* rare-variants and MD suggests our analysis is underpowered, or that genetic effects on MD are conferred substantially by higher frequency variants. Rare-variant MR suggested a causal effect of DHA on MD, and implicates the *FADS2* gene.

## Introduction

Major Depression (MD) is one of the commonest psychiatric disorders and is a leading cause of disability worldwide (1, 2). Researchers have identified a significant biological component to individual MD risk; the largest Genome wide association study (GWAS) identified nearly 700 common genetic variants associated with MD (3–7). However, the mechanisms through which genetics contribute to disorder risk remains poorly understood. A better understanding of these mechanisms would allow for improved predictive tools and drug development to treat MD.

Evidence is accumulating which suggests that various metabolic functions influence MD (8, 9). For example, two large Dutch cohort studies identified associations between depression and 21 and 79 metabolites respectively, measured in the blood (10, 11). Similarly a large-cross cohort study found predictors derived from metabolites outperformed genetic risk scores in predicting a range of diseases including depression (12).

Studies of diet and nutrition have also implicated metabolites in mental health. Correlations have been observed between the intake of long-chain polyunsaturated fatty acids (LU-PUFAs) such as Omega-3, on MD and other psychiatric disorders (13–15). LU-PUFAs are a family of molecules consisting of acids with long chains of double bonded carbons. They are primarily acquired through diet but are also synthesised in the body from shorter chain fatty acids (13). They have a range of psycho-active effects and along with derived molecules, have been implicated in neurogenesis, brain inflammation and synaptic function (16, 17).

Large-cohort genetic studies have found evidence for causal effects of LU-PUFAs in depression. A study by Davyson et. al in a sample of ∼30,000 individuals identified 129 metabolites measured in the blood robustly associated with MD; they found evidence of a putative causal role of 5 LU-PUFA measures on MD (18). Notably, these findings were dependent on variants in the *FADS* gene cluster on chromosome 11(18, 19). This region plays an important role in metabolism, specifically, in catalysing stages of the desaturation of LU-PUFA during biosynthesis (20). A similar Mendelian Randomisation (MR) study found causal variants in the *FADS* region effected multiple psychiatric disorders via 29 metabolite measures, implying a broader influence of the *FADS* region on psychiatric traits (21). In addition to metabolites, genetic studies of this region have implicated other molecular phenotypes in MD including gene expression and DNA methylation (3, 20, 22, 23). An improved understanding of causal mechanisms of the *FADS* region on MD could help guide nutritional advice and identify drug targets for future pharmacotherapies.

Most population genetic research has studied common variants, in which the minor (rarer) allele occurs in at least 5-10% of the studied population. However, GWASs of MD have found common-variants explain only 8-14% of the heritability of MD (compared to a total estimated heritability of ∼37%) (3, 24, 25). An additional limitation is that common variants in the *FADS* region are highly correlated with each other (in Linkage Disequilibrium/LD). Significant common variants may simply be correlated with truly causal variants making it challenging to identify the specific genes underlying these associations. Multiple genes in the cluster including *FADS1*, *FADS2*, *FADS3*, *MYRF* and *TMEM258* have plausible mechanisms through which they could affect MD (20).

Studying rare genetic variants in which the minor allele occurs in less than 1% of the population can potentially address these limitations. As large, whole genome-sequenced cohorts are becoming available, association studies of rare variants are becoming feasible and in MD, studies have found they contribute to heritability additively with common variants (26). Rare variants show far lower LD than common variants so causal effects can be mapped to specific genes in with greater confidence (27). Additionally, several studies have suggested that rare variant-trait associations map to the same genes and genomic regions as common variants (28–30). Variants with a large negative effect on gene function are also more likely to be rare, as negative selection prevents pathogenic variants becoming common in the population (31–33). Consequently, rare variants have significant potential to provide novel insights and identify the causal mechanisms of MD which cannot be elucidated using common-variant approaches.

To test this approach, we selected the *FADS* region for rare-variant analyses, as it has been previously implicated in MD by GWAS and metabolic studies, but without a clear causal gene identified. We aimed to identify variants and specific genes which effected MD risk via metabolic function.

Our study takes advantage of recently available, large-scale whole genome sequencing data in the UK Biobank (N = 466,864) and All of Us (N = 294,820) cohorts to test for associations of rare genetic variants on metabolites levels and MD. Using short-read whole genome sequencing data we tested for associations between rare variants in the *FADS* region, and MD, using both single-variant and aggregate testing methods. Summary statistics from both UKB and All of Us were meta-analysed for our final analysis. Additionally, we conducted rare-variant analysis on the 5 metabolites implicated in MD by Davyson et. al (18). Finally we conducted two-sample MR to determine if rare variant instruments can be used to identify causal effects of metabolite levels on MD.

## Methods

### Cohorts

We used phenotypic and genetic data from All of Us (AoU) and the UK Biobank (UKB), and Nuclear Magnetic Resonance (NMR) metabolomic data from UKB (34–36).

AoU is an American research program formed by the National Institute of health. The program aims to collect and study data from 1 million people age 18 and above people living in the USA for health research (N = 633,534 at the time of our analysis). For our analysis we selected only individuals for whom short-read Whole Genome Sequencing (srWGS) data, electronic health records (EHRs), age and sex were available.

UKB is a large prospective study tracking the long-term health of people as they age (N = 502,492). Participants were recruited between ages 40 and 69 from the United Kingdom. We selected individuals with an MD phenotype available (see below) and with available age, sex and srWGS data. The metabolite analysis was further restricted to those for whom metabolite data was available.

For full cohort details see the supplementary methods.

### Major Depression

In AoU, the MD phenotype was defined using EHRs. Individuals with at least one diagnosis of Major Depression recorded in their EHRs from a specialist healthcare provider were defined as cases (Concept ID 4152280). Individuals with a recorded diagnosis of schizophrenia (Concept ID 435783) or bipolar disorder were excluded (Concept ID 436665).

In the UKB, we used the MD phenotype previously used for the most recent MD GWAS from the Psychiatric Genomics Consortium (PGC) (3). For details see the supplementary methods.

### Metabolomic data

Metabolomic and srWGS data in UKB was available for 469,738 participants. We selected metabolites based on the results of Davyson et. al (2023) (18); specifically the five measures identified via Mendelian randomisation as being associated with MD using instruments in the *FADS* region: Docosahexaenoic Acid (Field ID: 23450), Omega-3 Fatty Acids (Field ID: 23444), Omega-3 to Total Fatty Acids (%) (Field ID: 23451), Omega-6: Omega-3 ratio (Field ID: 23459) and the Degree of Unsaturation (Field ID: 23443).

To remove technical variation in the metabolite data we used the ‘ukbnmr’ R package (37). Following QC, we performed rank-based inverse normal transformation on all metabolite data. Metabolite analysis was conducted in European ancestry only.

For details see the supplementary materials

### Genetic Variant extraction and QC

We used srWGS and microarray data from AoU and the UKB.

In both cohorts we selected rare variants from srWGS data in *FADS* gene cluster on chromosome 11 defined as chr11:61750000-chr11:61900000, which contains the following genes; *FADS1*, *FADS2*, *FADS3*, *FEN1*, *MYRF*, and *TMEM258* (Figure 1). srWGS variants were filtered to those with a MAF < 0.01.

**Figure 1.**
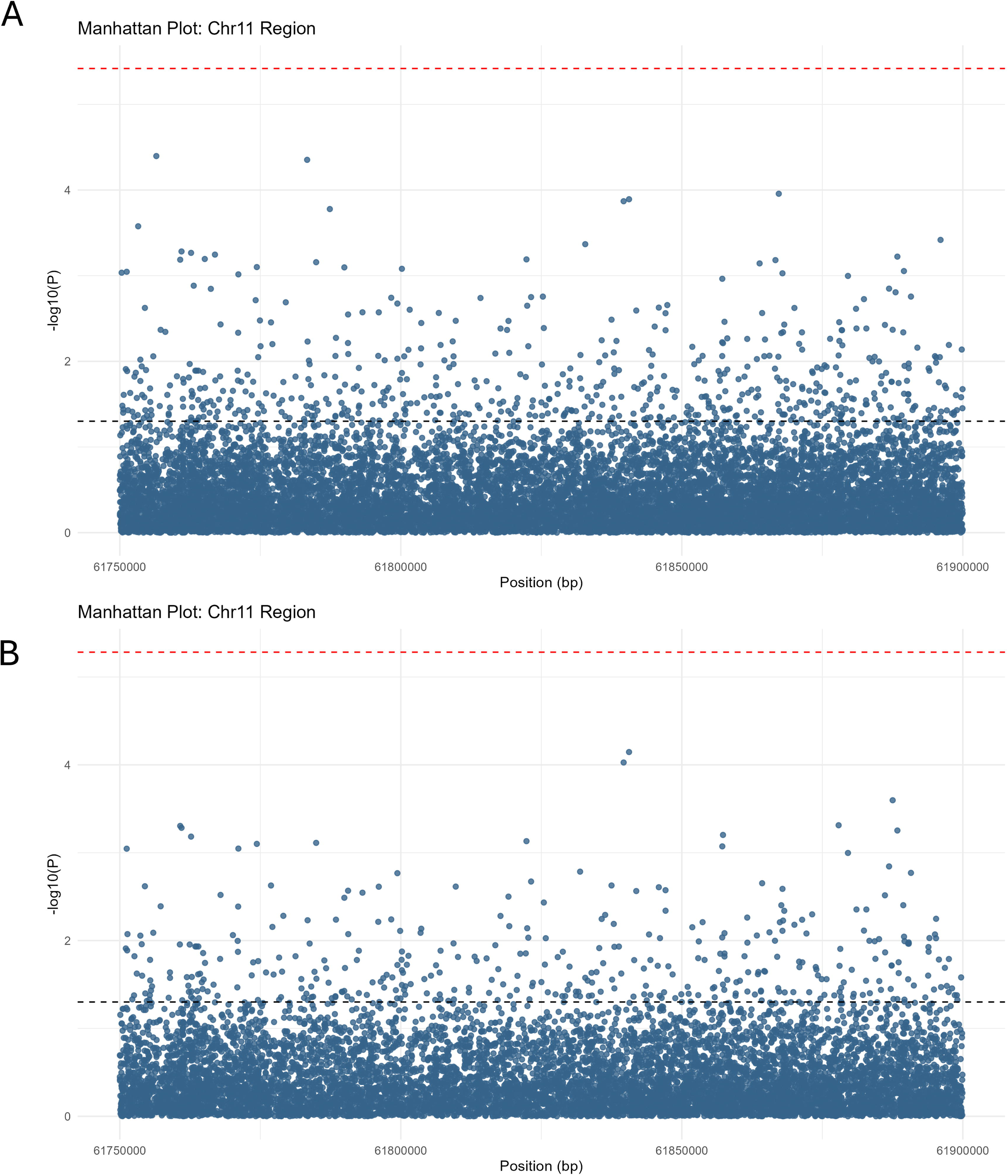
A visualisation of the FADS region. Strand is indicated by colour. Gene regions are shown as defined by the Ensembl database (version 115) (40)

In the UKB biobank we selected and concatenated the pVCFs containing srWGS data for this region into a single file with ‘bcftools’ version 1.18. In AoU, variants in this region were extracted from the VDS file and exported to sharded VCFs using HAIL version 0.2 (https://github.com/hail-is/hail). For both datasets, initial filtering and QC were conducted on the VCF files using ‘bcftools’ (38), and were subsequently converted to PLINK BED format (39) for further QC and analysis. For full details on rare-variant QC and microarray QC and use see the supplementary methods.

### Variant masks and annotation

Following variant filtering and QC, variants were annotated using the Ensembl Variant Effect Predictor (VEP) (Version 113, build GRCh38) (41). This uses the Sequence Ontology database (42) to categorise variants predicted impact on gene function as high, moderate, low and modifier.

Examples of high-risk variants include frameshift variants while examples of low and modifier risk variants included intronic and intergenic variants (https://www.ensembl.org/info/genome/variation/prediction/predicted_data.html). We classified variants by VEP Impact rating, taking the highest impact annotation for each variant, across all transcripts.

For aggregate testing variants were grouped into ‘Masks’, based on the functional annotations. Masks were defined using progressively more inclusive annotation criteria. Mask 1 included high impact VEP variants only, Mask 2 included High and Moderate impact, Mask 3 included High, Moderate, and Low, and Mask 4 included High, Moderate, Low and Modifiers (Table 1).

**Table 1.**
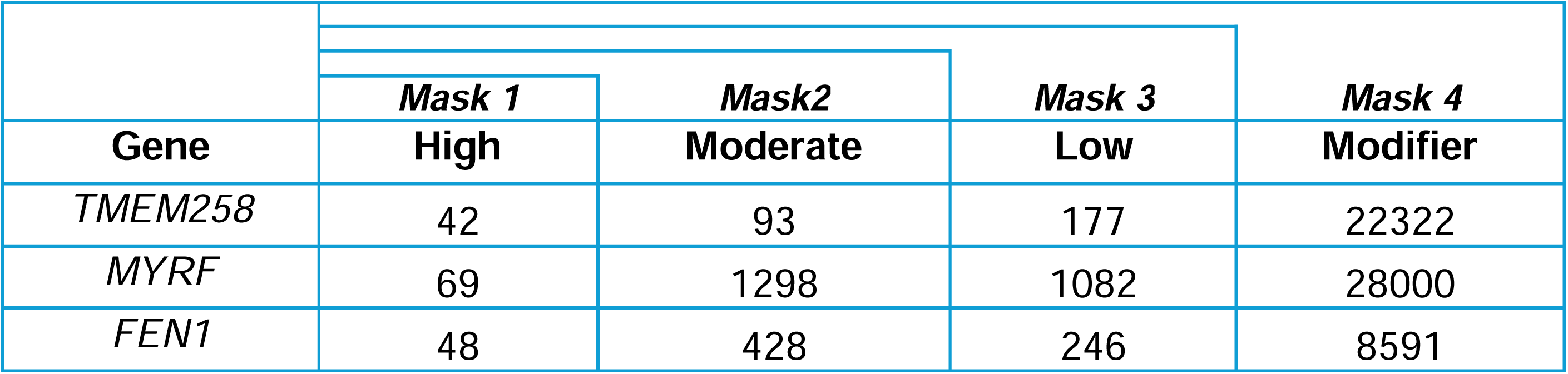
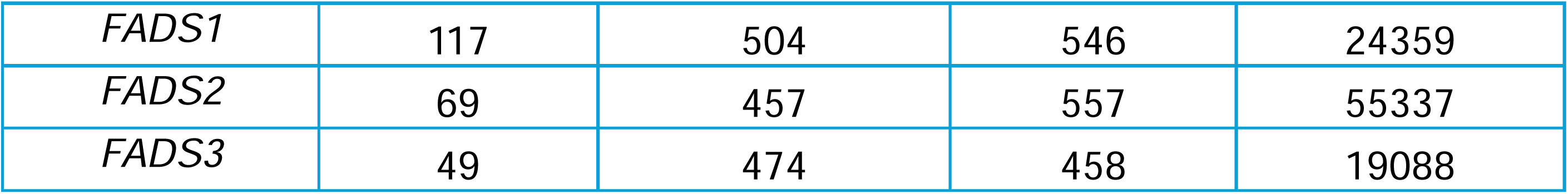
A breakdown of annotations by gene across both UKB and AoU, grouped by impact as defined by the Sequence Ontology database. Note: A single variant could have multiple annotations across different genes.

Separate masks following the same criteria were created for singleton variants.

### Rare variant association testing

We ran association tests between MD and rare variants, seperately UKB and AoU using REGENIE (version 4.1) then meta-analysed them using REMETA (version 1.1). In AoU tests were conducted in individual ancestries then meta-analysed (PCs were calculated seperately for each ancestry group).

We ran association tests between metabolite measures and rare variants in UKB only using REGENIE.

For a breakdown of how ancestries were combined in the meta-analysis and how covariates were calculated see the supplementary methods.

### Single variant association tests

For single-variant tests step 1 was run using genome-wide microarray data (MAF > 0.01), while step 2 was conducted on srWGS data filtered to rare-variants in the *FADS* region (maxMAF = 0.01). Variants with a minor allele count below 10 were excluded from the analysis. We applied a Bonferroni correction across all variant tests within each phenotype.

We included sex, age, the first 10 ancestry-specific genetic PCs as covariates and MD-PRS (MD analyses only). In UKB, we additionally included WGS batch, and in AoU we included sample source and data collection location.

Summary statistics from both cohorts were then meta-analysed using REMETA’s effect size meta-analysis, in European individuals only and across all-ancestries (Box S1) (43).

#### Aggregate tests

We ran gene-level aggregate testing across all QC’d variants using REGENIE in each of the 6 genes in the region. Aggregate testing groups variants into a single risk score to test their collective effect on a phenotype. As with the individual variant associations with step one used array-type data and step 2 used srWGS data of the *FADS* region (maxMAF = 0.01). We ran several types of aggregate test: Burden, SKAT, ACAT-V, and SKAT-O and ACAT-O Omnibus tests. For full details on the aggregate tests used see the supplementary methods.

Association tests were conducted separately in ancestry groups before meta-analysis using REMETA’s gene-based meta-analysis, in all ancestries and in Europeans only. Burden testing for the 5 metabolites of interest was conducted in UKB only. The meta-analysis did not include the ACAT-O tests as these were not available via REMETA.

All aggregate tests used the same mask definitions, tests and thresholds. In addition, they used the same covariates as the single-variant association analysis. We applied a Bonferroni correction across all tests and genes within each phenotype.

#### Leave-One-Variant-Out (LOVO) analysis on metabolites

Following burden testing we conducted LOVO analysis for all 5 metabolites using REGENIE. LOVO analysis re-runs aggregate testing on a mask multiple times, each time leaving out a single variant. Substantial changes in the p-value can then be used to identify variants with outside impacts on the effect of each mask on the phenotype.

LOVO tests were run on significant tests in Mask 3 for all metabolites as these also contained all the variants in Masks 1 and 2. Analysis was run using the burden, SKAT, SKAT-O, and ACAT-O tests.

For each gene we identified the variants which, when excluded, resulted in the greatest drop in the -log10 p-value and annotated them using the Ensembl VEP.

### Two Sample Mendelian Randomisation (MR)

We conducted Two sample MR using the ‘TwoSampleMR’ R package (19). We filtered variants to those with genome-wide-significant associations with metabolites, and which were present in both the UKB and All of Us sample. Metabolite levels in the UKB were treated as the exposure and MD in All of Us was treated as the outcome. The analysis was conducted using weights obtained from the European-only analyses in order to account for differing demographics across the cohorts. Variants were tested individually using Wald-ratio tests, and collectively per-metabolite using MR Egger, Weighted Median and Inverse Variant Weighted (IVW).

We filtered instrument variants to those significantly associated with a metabolite at Bonferroni-corrected test-wide significance, with a minor allele count > 10 and which was measured in both datasets. We applied Bejamini-Hochberg FDR correction across all tests within each metabolite.

### Post-hoc sensitivity analysis on statistical power

For single-variant associations with MD and Omega 3 (selected as a representative metabolite measure), we calculated the power to correctly reject null hypothesis for each tested variant given our parameters of sample size, case control ratio and a test wide significance threshold. We repeated using chromosome 11 of the largest GWAS of MD to date as a comparison (NCase = 688,808, NControl = 4,364,225, p < 5 x10^-8^). (3). We plotted the thresholds of the MAF and sample size required for a single variant association test to reach a power of 0.2, 0.5 and 0.8. We assumed the maximum number of variants was measured in all samples.

We calculated the percentage of tests in our analyses which reached a theoretical power of 0.8. Additionally we plotted the estimated sample size required for an individual variant test to be significant based on the median MAF and absolute median log(OR), using the same MD case control ratio as in our analysis.

For full details see the supplementary methods.

## Results

### Sample

For the MD analysis, following QC we retained 294,820 individuals (nCaseMD = 44,628, nControlMD = 250,192) in AoU and 466,864 individuals (nCaseMD = 69,691, nControlMD = 397,173) in UKB for whom genetic, phenotypic and covariate data was available across all ancestries. In the European only subset we retained 168,893 individuals (nCaseMD = 28,038, nControlMD = 140,855) in AoU and 445,553 individuals (nCaseMD = 67,460, nControlMD = 378,093) in UKB for whom genetic, phenotypic and covariate data was available.

For the metabolite analysis following QC we retained 448,224 individuals of European ancestry and for whom genetic, phenotypic and covariate data was available.

### Variant selection

Following QC we selected 57,915 variants UKB and 61,427 in AoU for analysis. There was an overlap of 26,364 rare variants between the two samples for a total of 100,221 variants across all ancestries. Variants were annotated using the VEP (Table 1). For a breakdown of annotations per-cohort see Figure S1.

### Rare variant associations with MD

#### Single variant association tests with MD

In our association tests between individual rare variants and MD we found no variants to be associated with MD at test-wide significance in either our all-ancestry or European only analysis (*pBonf* < 0.05). In the all-ancestry sample 618 variants were nominally associated with MD and, in the European-only sample 405 variants were nominally associated with MD (*p* < 0.05) (Figure 2).

**Figure 2.**
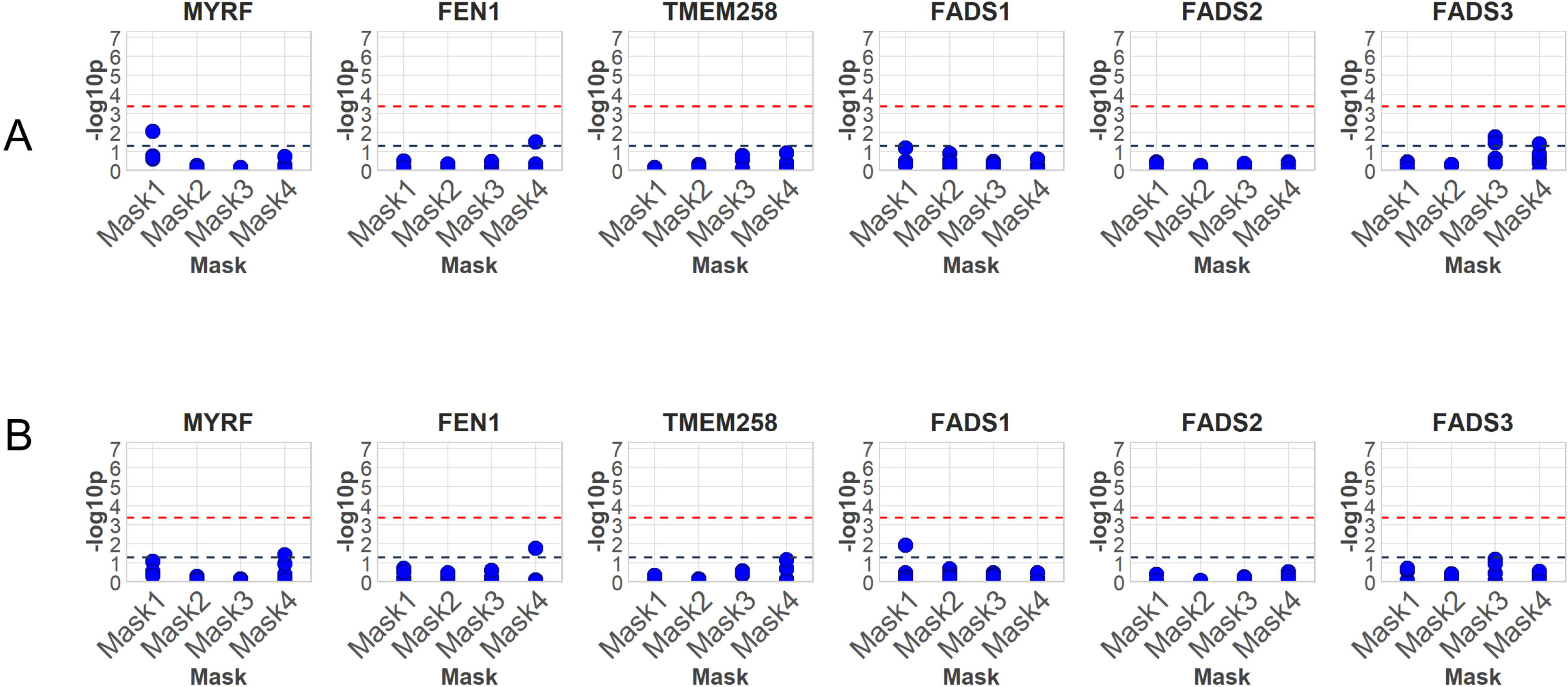
Manhattan plots for meta-analysed associations between individual rare variants and MD in A) all-ancestries and B) Europeans only. The black line indicates the nominal significant threshold (p < 0.05) while the red line indicates the Bonferroni-adjusted significance threshold.

#### Aggregate testing for MD

In our aggregate testing meta-analysis, no masks were significantly associated with MD following multiple testing correction in either the mixed ancestry or European only analysis (*pBonf* < 0.05) (Figure 3, Table S1-2). The top mask in the mixed ancestry analysis was the singletons mask in *MYRF* (*p* = 0.008). In the European only ancestry, the top mask was the singletons mask in *FADS1* (*p* = 0.012).

**Figure 3.**
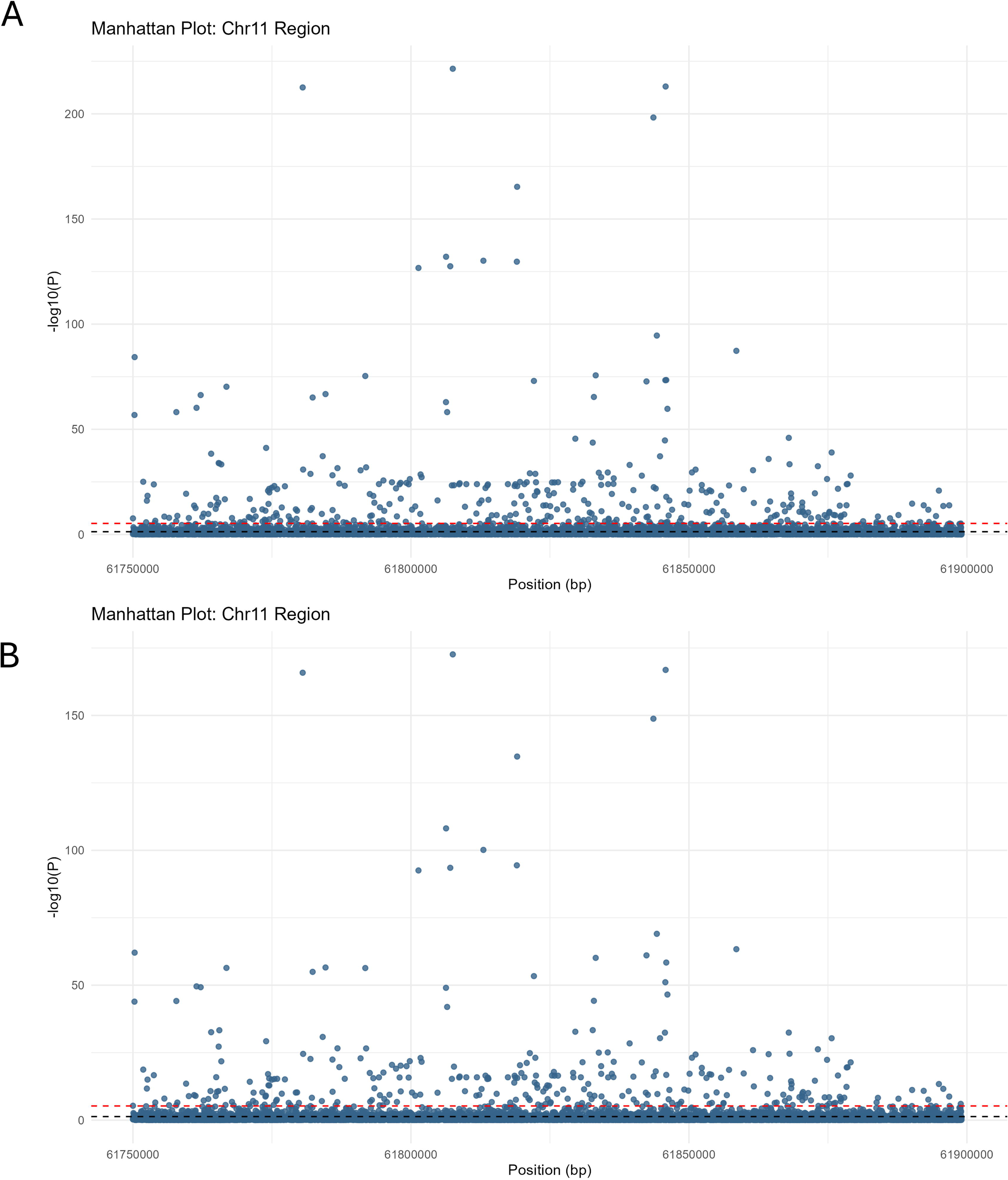
Plots depicting association between the burden of rare variants and major depressive disorder across different masks in A) all ancestries and B) Europeans only. The different types of tests for each mask (e.g. burden, SKAT etc) are represented by the multiple dots within each mask. The black line indicates nominal significance, while the red line indicates test-wide significance.

### Rare variant associations with Metabolites

#### Single variant association tests for metabolites

We identified a large number of variants which were associated with metabolite levels at test-wide wide significance (*pBonf* < 0.05): 303 with DHA, 357 with Omega 3, 441 with Omega 3 percentage, 417 with Omega 6 to Omega 3 ratio and 372 with Unsaturation (Figure 4). 254 variants were significantly associated with all 5 of our metabolite measures *pBonf* < 0.05), consistent with the high degrees of correlation between the measures (Figure S2, Table S3)

**Figure 4.**
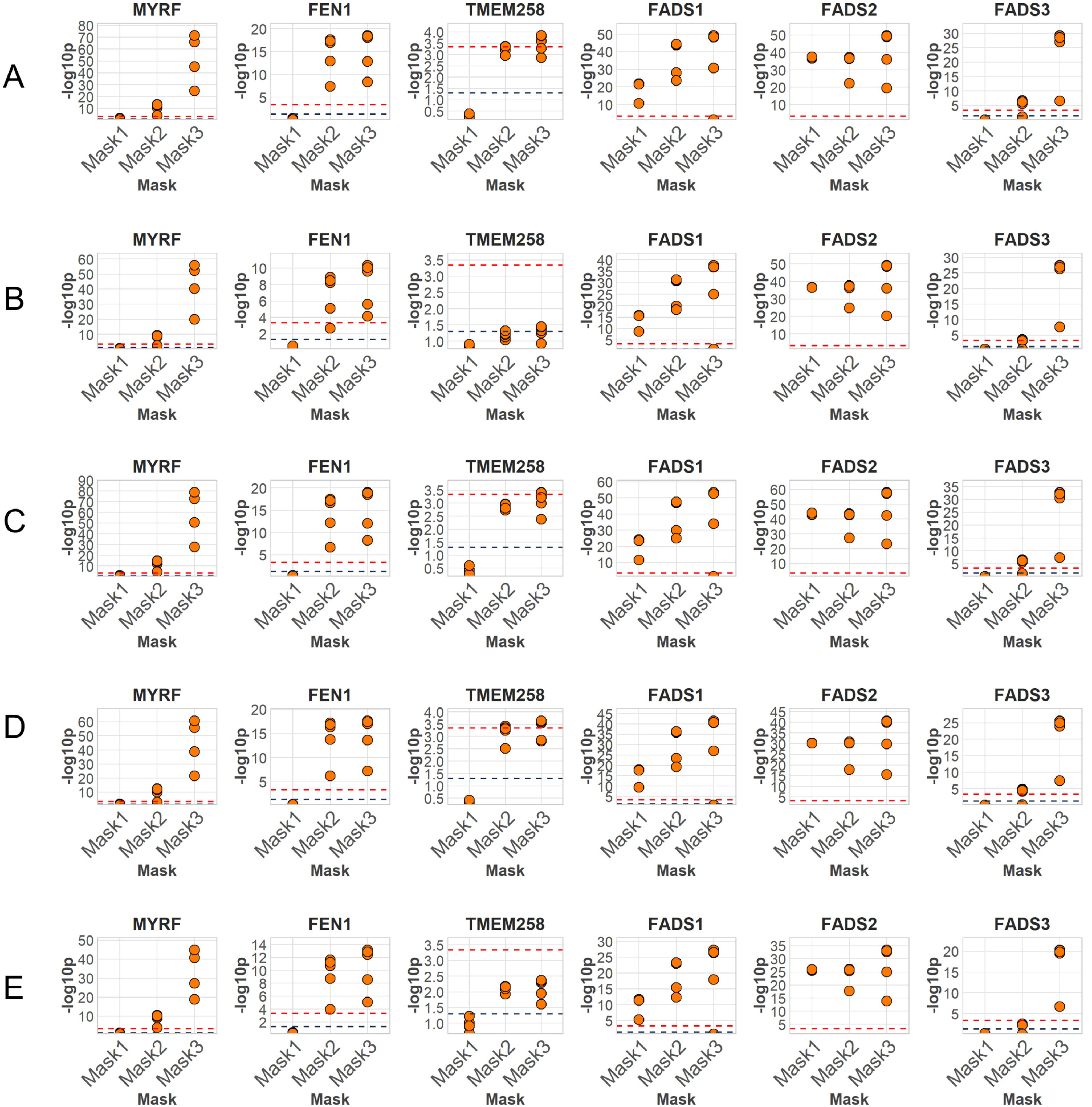
Manhattan plots for single variant associations with A Omega 3, and B) DHA. The black line indicates nominal significant threshold (p < 0.05) while the red line indicates the test-wide Bonferroni-adjusted significance threshold. For Manhattan plots for all metabolites see Figure S3.

#### Aggregate testing for Metabolites

Aggregate testing of metabolites identified widespread associations between rare variant burden and all metabolites in our analysis (*pBonf* < 0.05) (Figure 5). Across the five tested metabolites, at least 1 mask was significant in all 5 genes. Mask 4 was the most significant mask in all genes and we observed that the significance of associations increased along with the number of variants in the mask. Mask 1 was significant in *FADS1* and *FADS2* only (Table S4-S8).

**Figure 5.**
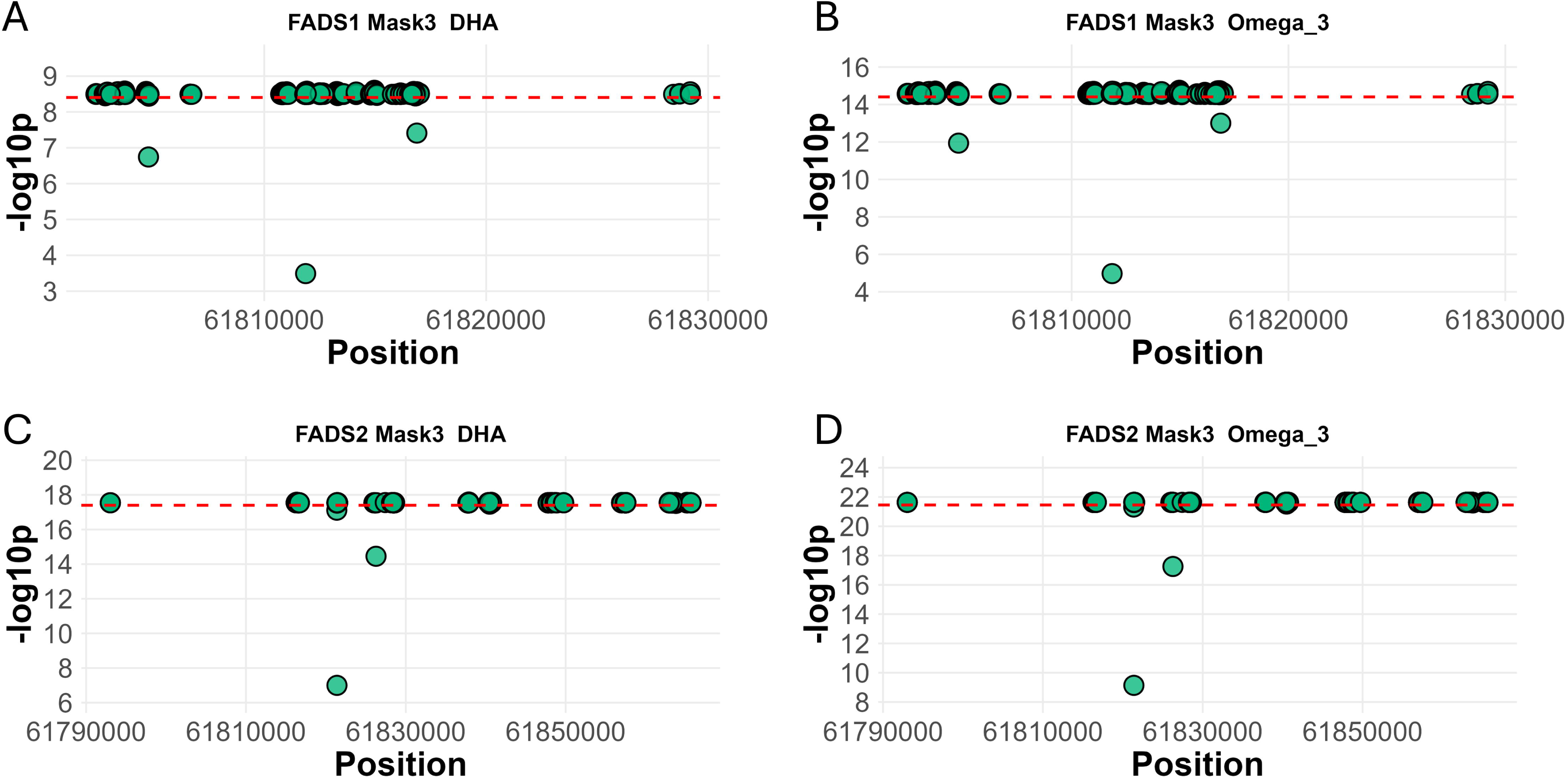
Plots depicting the associations between the burden of rare variants and metabolite levels across different masks in all ancestries. The black line indicates nominal significance, while the red line indicates test-wide significance. Note that the -log10p-values for Mask 4 were generally orders of magnitude greater than the in Masks 1-3. and so have been omitted for legibility. For plots of all four masks see Figure S4.

#### LOVO analysis

In the LOVO analysis on Mask 3, we found that the variants contributing most significantly to the burden tests for each gene was the same across all of our tested metabolites (Figure 6, Table 2).

**Figure 6.**
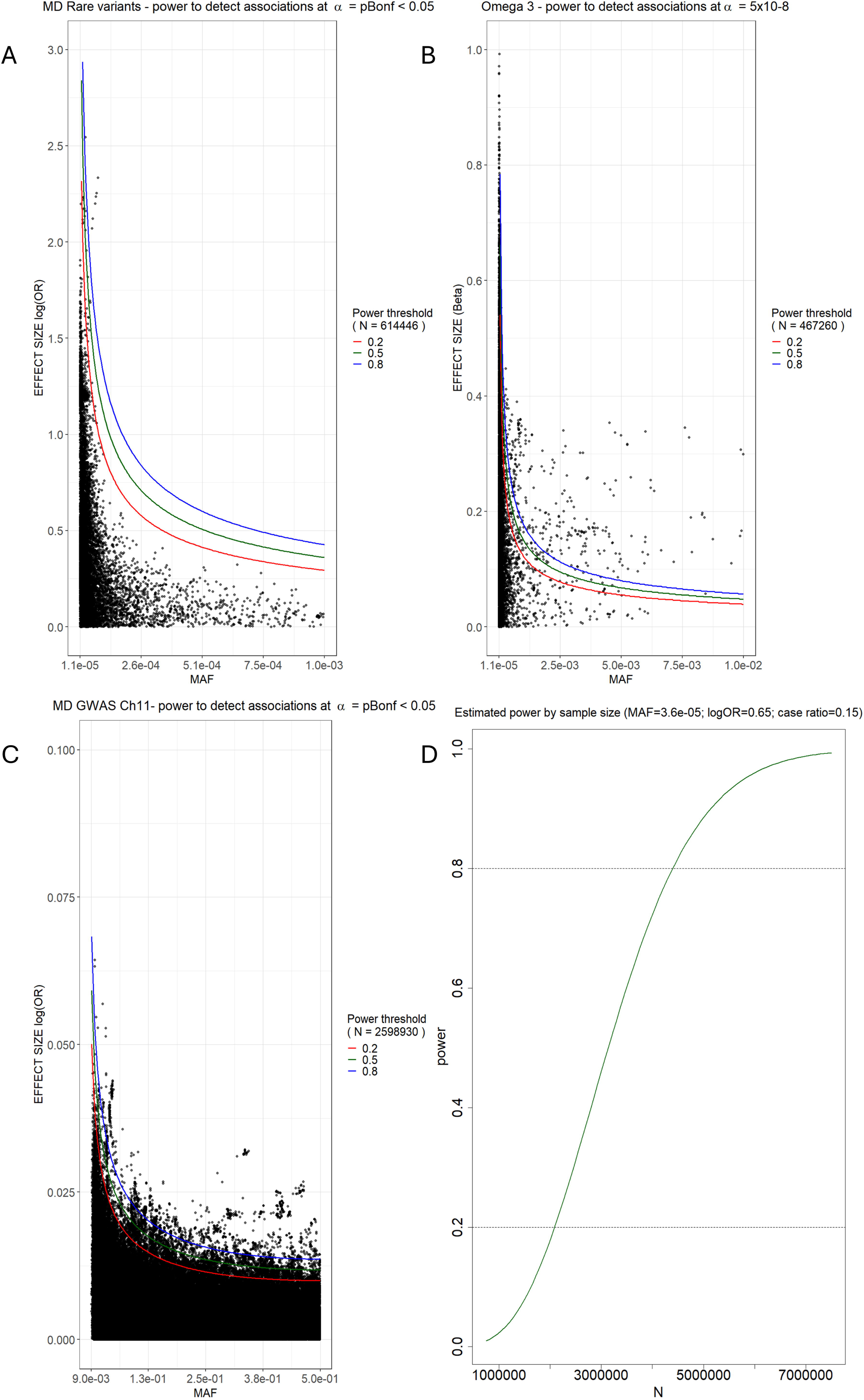
Plots displaying LOVO results between Mask 1 using the SKAT test in A) FADS1 on DHA, B) FADS 1 on Omega 3, C) FADS2 on DHA and D) FADS2 on Omega 3. Each dot represents test run for the specified mask and gene with a single variant removed. Decreases in - log10p-value indicate a variant contributing significantly to the overall burden of rare variants on the phenotype. The red line indicates the -log10p value of the test with all variants included.

**Table 2.** Table of the top LOVO identified variants contributing to significant rare-variant burden associations with metabolites. Each metabolite has the same top variant. The top variants for each gene as defined by influence on p-value and the accompanying VEP annotations. N.B. only the most pathogenic annotations have been reported for each variant.

| Gene | Variant | Impact | Predicted consequence (VEP) | MAF (gnomAD) |
| --- | --- | --- | --- | --- |
| *MYRF* | 11:61773957:G:G<br>C | Low | Splice polypyrimidine tract variant | 0.00285558 |
| *TMEM258* | 11:61789892:C:T | Moderate | Missense variant, NMD transcript variant | 0.00035277 |
| *FEN1* | 11:61796138:C:G | Modifier | Upstream gene variant | 0.00080851 |
| *FADS1* | 11:61811866:G:A | Moderate | Missense variant, Nonsense-mediated mRNA Decay (NMD) transcript variant | 0.00157302 |
| *FADS2* | 11:61821385:G:G<br>A | High | Frameshift variant | 0.00343321 |
| *FADS3* | 11:61878532:G:A | Low | Synonymous variant | 0.00361669 |

### Two Sample Mendelian randomisation

Across the five metabolites, we conducted between 277, and 391 Wald ratio tests. Following FDR correction, we found DHA was significantly associated with MD using chr11:61855776:C:T/ rs145534582 as an instrument (Beta = -2.250, SE = 0.597, p*FDR* = 0.046) rs145534582 was annotated by the VEP to *FADS2*, as an intronic variant to several transcripts. The MAF of this variant was 0.0018 in our sample.

We also applied multi-variant MR methods across each metabolite, finding no nominally significant associations (*p* < 0.05).

### Post-hoc sensitivity analysis

In our analysis of MD, we calculated that the discovery power of 0.8 was achieved in 0.06% of single variant tests (MAC > 10). For Omega 3 a power of 0.8 was achieved in 2.20% of single variant tests. For comparison, across chromosome 11 in the largest common-variant GWAS of MD, 0.68% of tests reached a power of 0.8.

Based on approximations of our median MAF (0.00002) and median beta (log(OR) = 0.68) we estimated that with our current case-control ratio, an association test would require a total sample size of approximately 4 million individuals in order to reach a discovery power of 0.8 (Figure 7).

**Figure 7.**
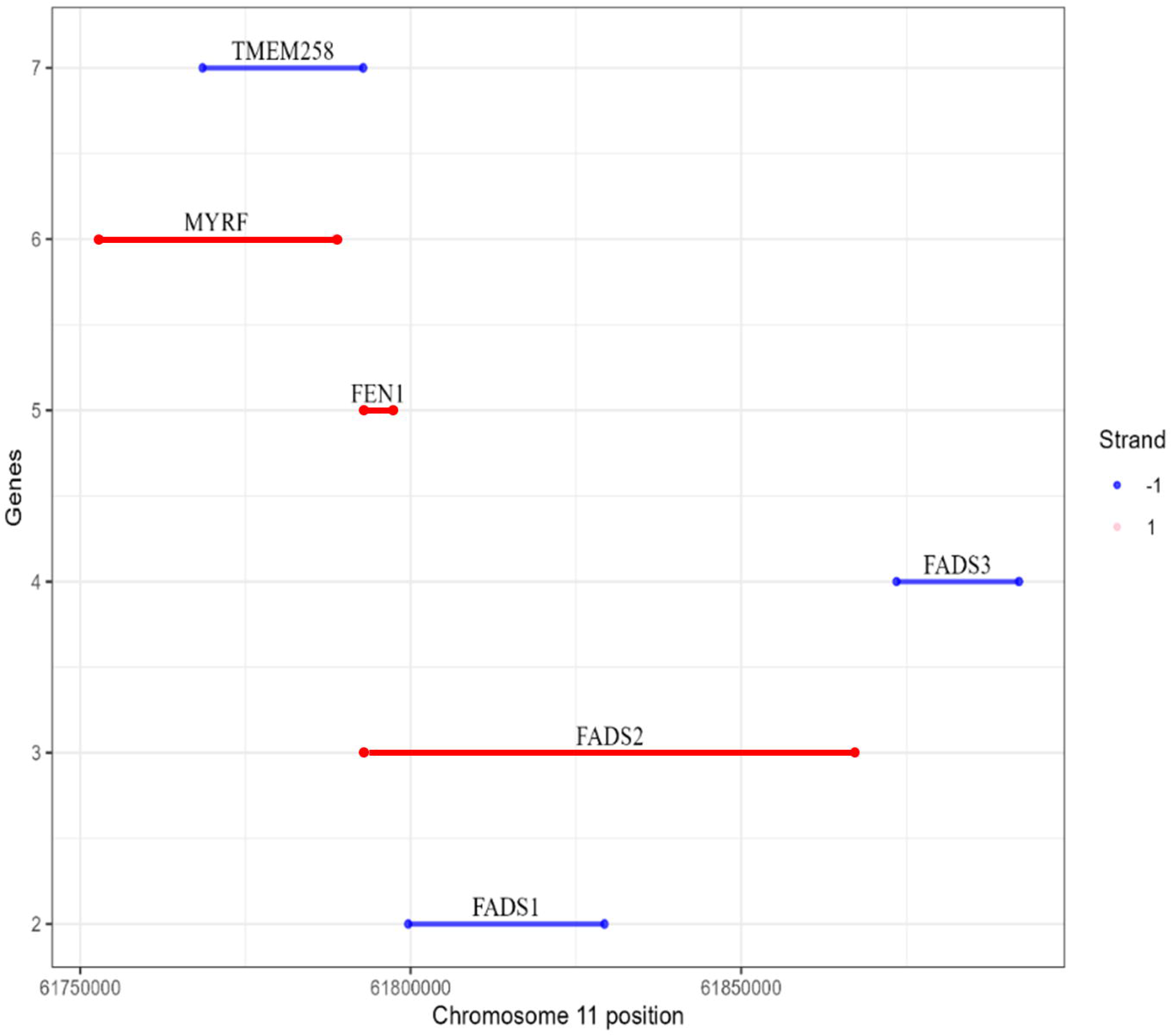
Plots showing the MAF/Effect size ratio of variants relative to the estimated discovery power, given known values of total sample size, case control ratio, and test-wide significance threshold in A) Rare variants in the FADS region tested for effects on MD, B) Rare variants in the FADS region tested for effects on Omega 3 and C) Common variants from chromosome 11 tested for effects on MD in the largest MD GWAS (3). D) Plot showing the estimated sample size required to reach a power of 0.8, for a variant with absolute median MAF (∼ 0.00002) and effect size (log(OR) 0.68), assuming a case control ratio of 0.15.

## Discussion

Our study analysed rare variants in the *FADS* gene cluster to better understand the mechanisms driving associations between metabolites and MD and identify causal genes. Rare variants in this region showed strong associations with all five metabolite measures previously associated with MD by Davyson et al. using common variants (18). However, we found no significant associations between rare variants in the region and MD. Applying Wald-ratio two-sample MR we observed a putative causal effect of DHA on MD, using a non-coding rare variant in *FADS2* as the instrument.

The null association between rare variants in the region and MD contrasts with previous research using common variants, both in GWAS and in molecular studies (3, 18, 21, 23, 44, 45).

Additionally rare-variant studies have found that variant burden was significantly associated with MD (26). However, they observed no significant associations in the *FADS* region; gene based testing identified only a handful of significant genes including *SLC2A1* and *NOG* in Tian et al. (26) and *OR8B4, TRAPPC11* and *SBK* in Cheng et al. (46)

Contrastingly, we observed significant effects of rare variants on metabolites, consistent with previous research implicating the *FADS* region in LU-PUFA regulation (47–49). Greater power to detect genetic associations with molecular phenotypes, relative to complex traits such as MD, has been repeatedly demonstrated; researchers attribute this to greater functional proximity between genetic variation and molecular endophenotypes (18, 50–54).

We observed a putatively causal effect of DHA on MD, aligning with the findings of Davyson et al. (18). The intronic instrument variant implicates *FADS2* in driving this effect; *FADS2* catalyses the first step LU-PUFA desaturation process, suggesting it is this initial step which is most relevant in metabolite influences on depression (55). Association studies have found substantial contributions of non-coding variants to the variance of both molecular and polygenic traits, including in depression (28, 29, 56). However the functional consequence of these variants remains poorly understood. Future work is needed to better understand the consequences of non-coding variants, and replication of our finding in independent samples will improve our confidence that the effect is truly causal.

Aggregate testing on MD, found that masks with more variants did not show lower p-values than smaller masks (Figure 3). Contrastingly, aggregate testing on metabolites showed a dosage effect, where burden test significance broadly increased in line with the number of variants included in the mask (Figure 5, Figure S4). One explanation is non-overlapping impacts between rare and common variants on MD; investigations into the convergence of common and rare variant effects have found that lab measures (such as metabolites), and anthropometric traits, show moderate to high degrees of convergence. However the effect is weaker in highly polygenic traits including depression, in line with our own results (29, 30, 46, 57). Therefore genetic effects on MD in the *FADS* region may be driven primarily by common variants.

Aggregate testing of metabolites showed similar patterns of significance across all five metabolites suggesting shared genetic effects across all measures (in line with Davyson et al. (18)). Additionally, neither single variant nor aggregate testing implicated a specific gene. However, among masks containing only high impact variants, only those for *FADS1* and *FADS2* were significantly associated with metabolites. This implies loss of function in these genes has a higher impact on metabolite measures than the other genes in our analysis. LOVO analysis identified one or two variants per mask which had a particularly large influence on association significance, the majority of which were high-impact variants (Table 2). This suggests both low and high impact variants contribute to metabolite regulation, with aggregate test associations being especially driven by high impact variants, in line with previous studies (30, 58, 59).

We conducted post-hoc analysis of our results and found our study to be generally underpowered to detect effects in MD, and may explain the lack of a variant-dosage effect in our aggregate tests. Power in GWAS is a function of allele frequency, effect size, sample size, significance threshold and, if applicable, case-control ratio (60, 61) (see Supplementary Methods). As allele frequency decreases, effect sizes and/or sample sizes of rare variants must increase to achieve comparative power to detect effects (62). However we found that even with a sample size exceeding 700,000, few tests were sufficiently powered to reject the null hypothesis compared to GWAS in common variants (Figure 7). We calculated that a tested variant with the absolute median values of MAF and effect size on MD that we observed in our analysis, would require a sample size exceeding 4 million to reach a power of 0.8 at test-wide significance (Figure 7).

A limitation to our analyses, is that they are restricted to a small section of the genome (63). While this targeted approach allowed for a detailed investigation of gene-specific rare variant effects and their relationship to relevant molecular endophenotypes, a data-driven search spanning the entire genome may provide additional insights into the broader genetic architecture of rare variants in MD disease risk. Another consideration is heterogeneity of MD phenotyping; to maximise sample size, we applied broad definition of MD. However, Tian et al. observed that association patterns with rare-variants differed depending on the definition of depression used (26). Our approach means the impact of rare-variant on depression heterogeneity is not captured (64).

In conclusion, our results suggest that we currently lack the sufficient sample size to replicate common variant associations in MD using rare-variants. This may also be due to limited regional overlap between common and rare-variant signals for MD. We did observe significant associations between rare-variants and metabolites, and found a putative causal effect of DHA on MD, in line with previous studies. We suggest that omic-data has the potential to be integrated with WGS to identify molecular associations with MD. Such analysis could have significant implications in understanding molecular pathways affecting complex traits.

## Data Availability

Data from both resources are available to authorized researchers subject to application and approval. Data produced in this study are available upon reasonable request.

## Acknowledgements

Individual level data analysis and processing was conducted on the UK Biobank (UKB) research Analysis Platform (https://ukbiobank.dnanexus.com) and the All of Us (AoU) Researcher workbench (https://workbench.researchallofus.org).

AMM and XS are supported by two UK Research and Innovation awards: Finding immune & metabolic pathways to SMI (Now ImmunoMIND) (Ref: MR/Z50354X/1), and the Hub For Metabolic Psychiatry (Ref: MR/Z503563/1)

AMM is also supported by a Wellcome Trust Investigator Award in Science (‘Exploiting genomic approaches to identify the environmental basis of depression’. (Reference: 220857/Z/20/Z), a UKRI award for the Hub for Metabolic Psychiatry (MR/Z503563/1), a UKRI award for the UKRI Mental Health Platform (MR/Z000548/1) and a European Union Horizon funding grant (CoMorMent: Predicting comorbid cardiovascular disease in individuals with mental disorder by decoding disease mechanisms” (Grant agreement 847776).

ED was supported by the United Kingdom Research and Innovation (grant EP/S02431X/1), UKRI Centre for Doctoral Training in Biomedical AI at the University of Edinburgh, School of Informatics.

With RB, the research was carried out at the National Institute for Health and Care Research (NIHR) Exeter Biomedical Research Centre (BRC).

Claude (Sonnet 5 and Opus 4.8) was used to review and provide commentary on human drafted text, to review code, and to generate code for the purpose of visualising data. No raw data was provided to the model and all outputs were reviewed by a human.

## Conflicts of interest

AMM reports research funding awarded by The Sackler Trust more than 5 years ago and PhD costs for LN were provided from funds previously awarded to the University of Edinburgh by The Sackler Trust more than 5 years ago.

At the time of submission ED is an employee of Genomics England.

All other authors report no biomedical financial interests or potential conflicts of interest.

## Notes

### Author Declarations

This research has been conducted using the UK Biobank Resource under Application Number 4844. UK Biobank has approval from the North West Multi-centre Research Ethics Committee as a Research Tissue Bank, and all participants provided written informed consent. The data are available from UK Biobank subject to application and approval The All of Us Research Program is supported by the National Institutes of Health, Office of the Director: Regional Medical Centers; Federally Qualified Health Centers; Data and Research Center; Biobank; The Participant Center; Participant Technology Systems Center; Communications and Engagement; and Community Partners.

### Summary of Updates

Added authors that the system removed

