## Supplementary Materials for "A rare-variant analysis of *FADS*-related metabolites in Major Depression"

Contents

1. *Supplementary Methods*
2. *Figure S1 – Per-cohort variant annotations*
3. *Figure S2 – Correlation heatmap of post QC Metabolite measures*
4. *Figure S3 – Manhattan Plots for single variant association tests on Metabolites*
5. *Figure S4 – Burden Test results for Metabolites Masks 1-4*
6. *Supplementary Table 1 – Aggregate testing results for MD (All ancestries)*
7. *Supplementary Table 2 – Aggregate testing results for MD (Europeans only)*
8. *Supplementary Table 3 – Rare variants associated with metabolites at test-wide significance*
9. *Supplementary Tables 4-9 – Aggregate testing results for metabolites*

Supplementary Methods

Cohorts

AoU is an American research program formed by the National Institute of health. The program aims to collect and study data from 1 million people living in the USA for health research (N = 633,534 at the time of our analysis). Participants are aged 18 and above, and data was collected across a wide range of health, lifestyle and genetic measures. Participants were selected from a diverse range of ethnicities, with 46% of participants being from underrepresented racial and ethnic minorities (1). Recruitment was conducted through a network of 22 community partners (2). Protocol, access and guidance procedures were approved by the AoU institutional review board following guidance from the Office for Human Research Protections. For our analysis we selected only individuals for whom short-read Whole Genome Sequencing (srWGS) data, electronic health records, age and sex were available.

UKB is a large prospective study tracking the long-term health of people as they age (N = 502,492). Participants were recruited between ages 40 and 69 from the United Kingdom. The cohort is predominantly white/European with over 90% of participants matching this description. Blood sampling and phenotype data collection was completed between 2006 and 2010. The North-West Research Ethics Committee reviewed and approved UKB’s scientific protocol and operational procedures (11/NW/0382). The analysis and data acquisition were conducted under application 4844. For our analysis we selected individuals with an MD phenotype (see below) available and with available age, sex and srWGS data. Likewise, the metabolite analysis was restricted to those for whom metabolic and srWGS data was available.

Short-Read Whole Genome Sequencing Data

In AoU srWGS data was available for 414,830 participants at the time of our analysis. Briefly, the AoU srWGS data processing was as follows: genomic data was extracted and sequenced across 4 Genome Centres (Baylor College of Medicine, Broad Institute and University of Washington (2018 – present) and the HudsonAlpha Institute of Biotechnology (2019 -present). WGS libraries were constructed with the Illumina Kapa HyperPrep kit, and were then pooled and sequenced on Illumina NovaSeq 6000 instruments, to a minimum coverage of 30x. Once collected, initial QC analysis was conducted with the Illumina DRAGEN pipeline, which include mapping, aligning, sorting, duplicate marking and haplotype variant calling. Genomes were aligned to the GRCh38 reference genome. All genome centres used harmonised laboratory protocols and DRAGEN parameters to maximise comparability (2).

In UKB we used the initial release of the DRAGEN WGS data generated by the DRAGEN v3.7.8 germline variant calling pipeline. SrWGS data was available for 490,549 participants, and was extracted and sequences via a public-private partnership between UK Biobank, UK Research and Innovation, Wellcome and four industry partners (Amgen, AstraZeneca, GSK and Johnson & Johnson). Sequencing was performed in two centres (deCODE facility in Reykjavik, Iceland and the Wellcome Sanger Institute (Sanger) in Cambridge, UK). Out of the 490,549 primary samples, 49,934 samples were sequenced as part of the Vanguard phase, 193,093 and 247,522 samples were sequenced by Sanger and deCODE, respectively. Sequencing was conducted on the Illumina NovaSeq 6000 platform, to an average coverage of 58x (and minimum of 10x). Processing was done using the Illumina DRAGEN Bio-IT Platform Germline Pipeline v3.7.8, and reads were aligned to the GRCH38 reference genome. QC criteria included coverage depth, genotype and mapping quality scores, DRAGEN variant status, alternate allele read proportion for heterozygous calls, proportion of samples failing any of these QC criteria, and gnomAD-related filters (3).

Genetic QC – srWGS and microarray

Variants were extracted into sharded VCFs from the VDS (AoU) and pVCFs (UKB), and combined using bcftools version 1.18. Once variants had been combined into a single file, calls were left-aligned and multi-allelic sites were split into biallelic variants, using the DRAGEN reference genome provided by the UKB (GRCh38_full_analysis_set_plus_decoy_hla.fa) and file (Homo_sapiens_assembly38.fasta) provided by AoU.

In AoU, variants were first filtered to remove those which failed the QC controls defined by AoU (i.e. those not labelled Filter = PASS in the VCFs). To generate this label, AoU conducted extensive pre-release quality checks as outlined in its genomic quality report (<https://support.researchallofus.org/hc/en-us/articles/29390274413716-All-of-Us-Genomic-Quality-Report>). Briefly, an initial sample level QC was applied, which involved comparing concordance between array and srWGS data, removing samples with low coverage, mismatches with reported and genetic sex, and outliers in metrics such as deletions and SNP count. Failing samples were removed from the data, Subsequently variant level QC was applied using hard filters for criteria including QUAL score, excess heterozygosity and excess alleles.

Once we had filtered by the hard thresholds defined by AoU (FILTER = PASS) we subsequently set any remaining genotypes with a Genotyping Quality (GQ) < 20 or sum of Allele Depth (AD) < 8 to missing. Lastly we removed any variants with a call rate < 0.9, a Hardy-Weinberg Equilibrium (HWE) p-value < 1x10-15 or which were monomorphic following QC.

We removed samples which failed AoU sample level QC, with missing genetically determined sex or mismatches between genetically determined sex and reported sex at birth and any samples with a variant call rate < 0.9. Lastly we calculated F coefficients for heterozygosity and filtered individuals with a coefficient > 3 standard deviations from the population mean to account for large structural variants in the region.

In UKB variants were likewise filtered to those that passed the default quality controls defined by the DRAGEN pipeline (<https://developer.illumina.com/dragen/dragen-popgen>). Allele frequencies for each variant were calculated from the samples. We set to missing any genotypes with GQ < 10 or, sum of local allele depth (AD/LAD) < 8. We then applied a variant level filter removing any variants with a call rate < 0.9, those with an HWE p-value < 1x10-15 or which were monomorphic following QC. Variant QC thresholds in UKB in line with previous rare-variant analyses (4, 5).

We then filtered samples to remove those with a mismatch between their genetically inferred and self-reported sex at birth, those with a sample level call rate < 0.9 and those with heterozygosity F statistics > 3 SDs from the population mean.

As our selected srWGS data covered only a small section of the genome, for REGENIE step 1, polygenic score calculation, and PCA calculation, we used genotype micrarray data. In the UKB we used array-based genotype data collected in 488,000 UK biobank participants by Affymetrix on the UK Biobank Axiom array. Genotype data was converted to GRCh38 and filtered to the participants included in the analysis. In AoU, we used the microarray data collected in 447,278 participants on the Illumina Global Diversity Array. In both UKB and AoU, we then filtered the common variants to remove any with MAF < 0.01 , HWE p-value < 1e-6, or those with a sample or genotype missingness rate > 0.1.

Metabolomic data

UKB Metabolomic data was collected and processed in partnership with Nightingale Health Plc. Metabolic profiling was conducted in EDTA plasma using Nightingale Health’s Nuclear Magnetic Resonance imaging (NMR) platform. The samples were prepared directly in 96 well-plates by the UK Biobank. Two NMR spectra were recorded for each plasma sample using a 500 MHz NMR spectrometer (Bruker AVANCE IIIHD). Automated quality control of the spectral data was performed. The metabolic biomarkers were quantified using Nightingale Health’s proprietary software (Nightingale Health biomarker quantification library 2020)(6)

To remove technical variation in the metabolite data we used the ‘ukbnmr’ R package (7). This package is custom made for UKB and corrects for a wide variety of technical variation in the metabolite data, including within plate structure, outliers and sample prep time. Following QC, we performed rank-based inverse normal transformation on all metabolite data. Metabolite analysis was conducted in European ancestry only.

Major Depression phenotype in UKB

In the UKB, we used the MD phenotype previously used for the most recent MD GWAS from the Psychiatric Genomics Consortium (PGC) (8). Under this definition patients meeting at least 2 of the following criteria were considered cases: Self-reported MD, self-declared anti-depressant treatment, at least one GP diagnosis of MD, three or more past GP prescriptions of antidepressions and meeting the threshold score for lifetime MD in the Composite International Diagnostic Interview (CIDI) questionnaire. In addition, participants with three or more GP diagnoses, or an inpatient diagnosis of MD recorded in EHRs were treated as cases. Individuals with a self-diagnosis or an EHR diagnosis of the following disorders were excluded: schizophrenia or schizotypal disorder, psychosis, mania, or bipolar disorder.

Calculating covariates for association analyses

In AoU we calculated the first 10 genetic principle components (PCs) for each ancestry group as defined by the genetically predicted ancestries in the AoU srWGS auxiliary filles using the microarray data. In UKB we calculated the first 10 PCs for European only and the all ancestry samples.

Briefly, in both cohorts, after removing related individuals and pruning to SNPs which were roughly independent (window = 50 SNPs, LD threshold r2 < 0.2) we calculated PCs then projected them into the full sample. Calculations in both cohorts used PLINK 1.9.

Polygenic scores in both cohorts were calculated using array data using SBayesRC using a European LD reference from the UK Biobank and annotation provided by SBayesRC (9). Weights were obtained from the largest available GWAS of Major Depression (8). For the UKB polygenic scores were calculated using weights calculated without the UKB sample.

Rare variant association testing

Association tests between MD and rare variants was conducted across both UKB and AoU. Association tests between metabolite measures and rare variants was conducted in UKB only.

For the association tests with MD, analyses in AoU were conducted separately in each genetically predicted ancestry group (European, Middle Eastern, African American, Admixed American, East Asian and South Asian) as defined by the srWGS auxiliary data prior to meta-analysis. Analysis in UKB was conducted in Europeans only and across all ancestries for meta-analysis. Genetic principal components were calculated individually for each ancestry specific analysis. Meta analysis for both single variant and aggregate testing was conducted using REMETA (version 0.9) (10). For a breakdown of how ancestries were combined see box 1. For a description of how covariates were calculated see the supplementary methods.

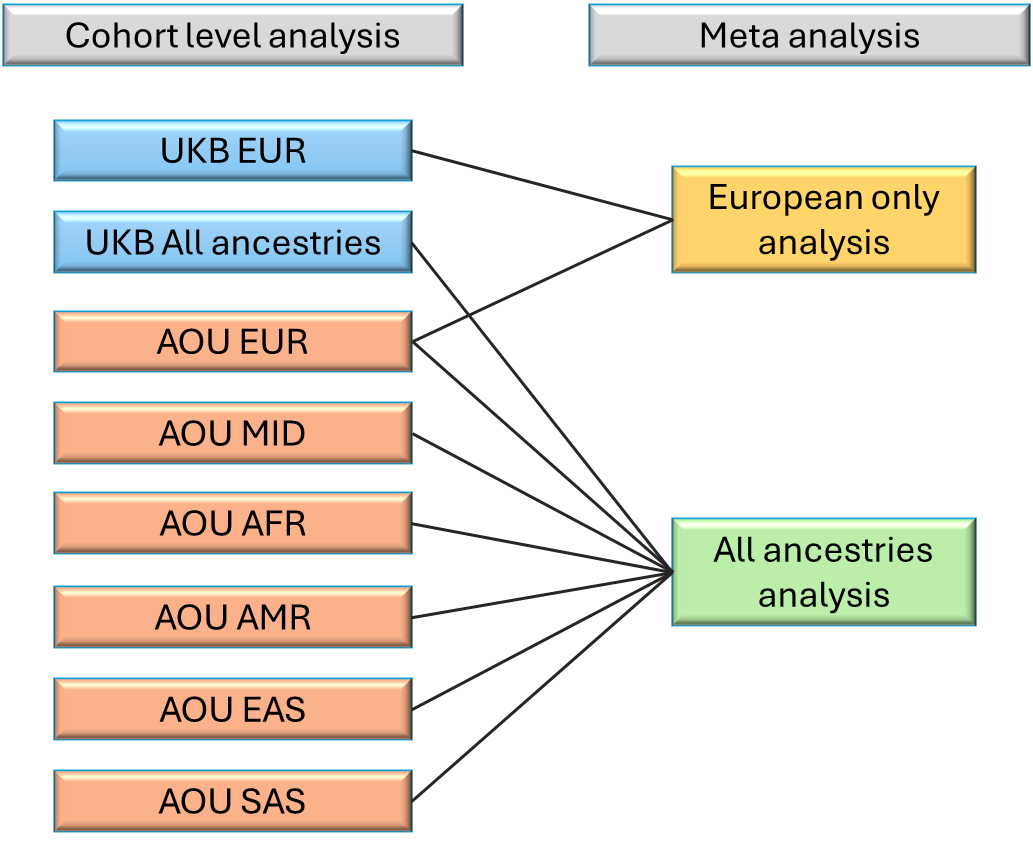

*Box 1. Flow chart indicating how different ancestry groups were combined for the final analysis of MD. Black lines indicate that the cohort was included in the meta-analysis with REMETA.*

Aggregate testing methods

There are various benefits and drawbacks, as well as assumptions, to different aggregate testing techniques, as such we implemented several methods in order to ensure comprehensive analysis. The four types of aggregate tests we implemented in REGENIE are: 1) burden test, which assume all variants in the mask have the same effect direction and roughly the same effect size, 2) sequence-kernel association test (SKAT), assumes that effect directions and magnitudes can vary between variants tested, 3) aggregated Cauchy combination test (ACAT-V) test, assumes varying effect direction varies across causal variants and adjusts for disparity in effect size or small number of associated variants, and 4) SKAT and ACAT-Omnibus (SKAT-O and ACAT-O) tests which adjusts the tests used to maximise statistical power, with minimal assumption on consistent direction of effects and is particularly suitable when a small number of variants are causal (4). Additionally, joint-burden tests (MIN_P, SBAT and GENE_P) were applied to test for associations across whole genes across all masks.

Calculating power in single variant tests

Power in variant tests is a function of sample size, MAF, effect size, alpha-threshold and case control-ratio (11, 12) . By inputting known values for sample size, case, control ratio, and alpha, we were able to plot power as a function of effect size and MAF and compare this the actual distribution of our single variant associations (MAC > 10). In addition estimates of power are less reliable in rare variants (13).

For MD (as a binary phenotype) we calculated the non-centrality parameter (NCP) of the additive model (under the assumption of independent variants) as :

$${NCP}_{bin}=(\beta/{SE}_{bin})^{2} \approx2f\left( 1-f \right)n{\phi(1-\phi)\beta}^{2}$$

And for metabolites (as a continuous phenotype) we calculated the NCP of the additive model (under the assumption of independent variants) as :

$${NCP}_{lin}=(\beta/{SE}_{lin})^{2} \approx2f\left( 1-f \right)n\beta^{2}/\sigma^{2}$$

$\beta$ = effect size (logOR in binary phenotypes, beta in continuous, $f$ = MAF, $n$ = total effective sample size, $\sigma^{2}$ = trait variance and $\phi$ = proportion of cases to controls) (14)

We plotted these as functions of MAF and Effect size and compared these to the variants in our tests (Figure 8)

Supplementary Figures

A

| UKB |  |  |  |  |
| --- | --- | --- | --- | --- |
|  | Mask 1 | Mask2 | Mask 3 | Mask 4 |
| **Gene** | **High** | **Moderate** | **Low** | **Modifier** |
| TMEM258 | 25 | 57 | 95 | 13155 |
| MYRF | 40 | 779 | 662 | 16894 |
| FEN1 | 24 | 250 | 125 | 5029 |
| FADS1 | 65 | 308 | 316 | 14274 |
| FADS2 | 38 | 283 | 337 | 32298 |
| FADS3 | 29 | 289 | 279 | 11242 |

B

| AOU |  |  |  |  |
| --- | --- | --- | --- | --- |
|  | Mask 1 | Mask2 | Mask 3 | Mask 4 |
| **Gene** | **High** | **Moderate** | **Low** | **Modifier** |
| TMEM258 | 28 | 64 | 136 | 15932 |
| MYRF | 50 | 904 | 780 | 19335 |
| FEN1 | 32 | 314 | 192 | 6092 |
| FADS1 | 74 | 318 | 382 | 17161 |
| FADS2 | 47 | 300 | 413 | 39428 |
| FADS3 | 24 | 322 | 341 | 13727 |

*Figure S1. Variant annotations in A) UK Biobank and B) All of us. For a breakdown of the categorisation criteria see refer to the Ensemble VEP guidance. (*[*https://www.ensembl.org/info/genome/variation/prediction/predicted_data.html*](https://www.ensembl.org/info/genome/variation/prediction/predicted_data.html)*)*

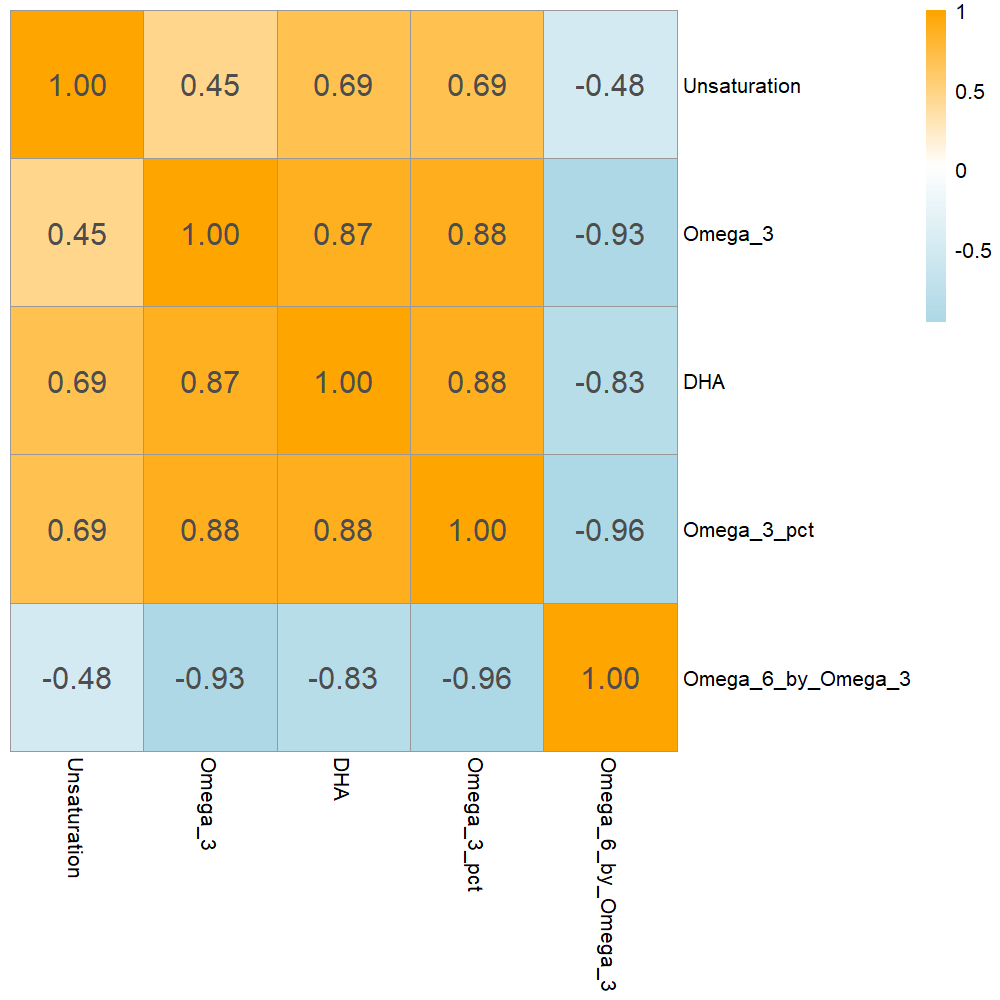

*Figure S2. Heatmap displaying the Pearsons correlation coefficients of the 5 metabolite measures.*

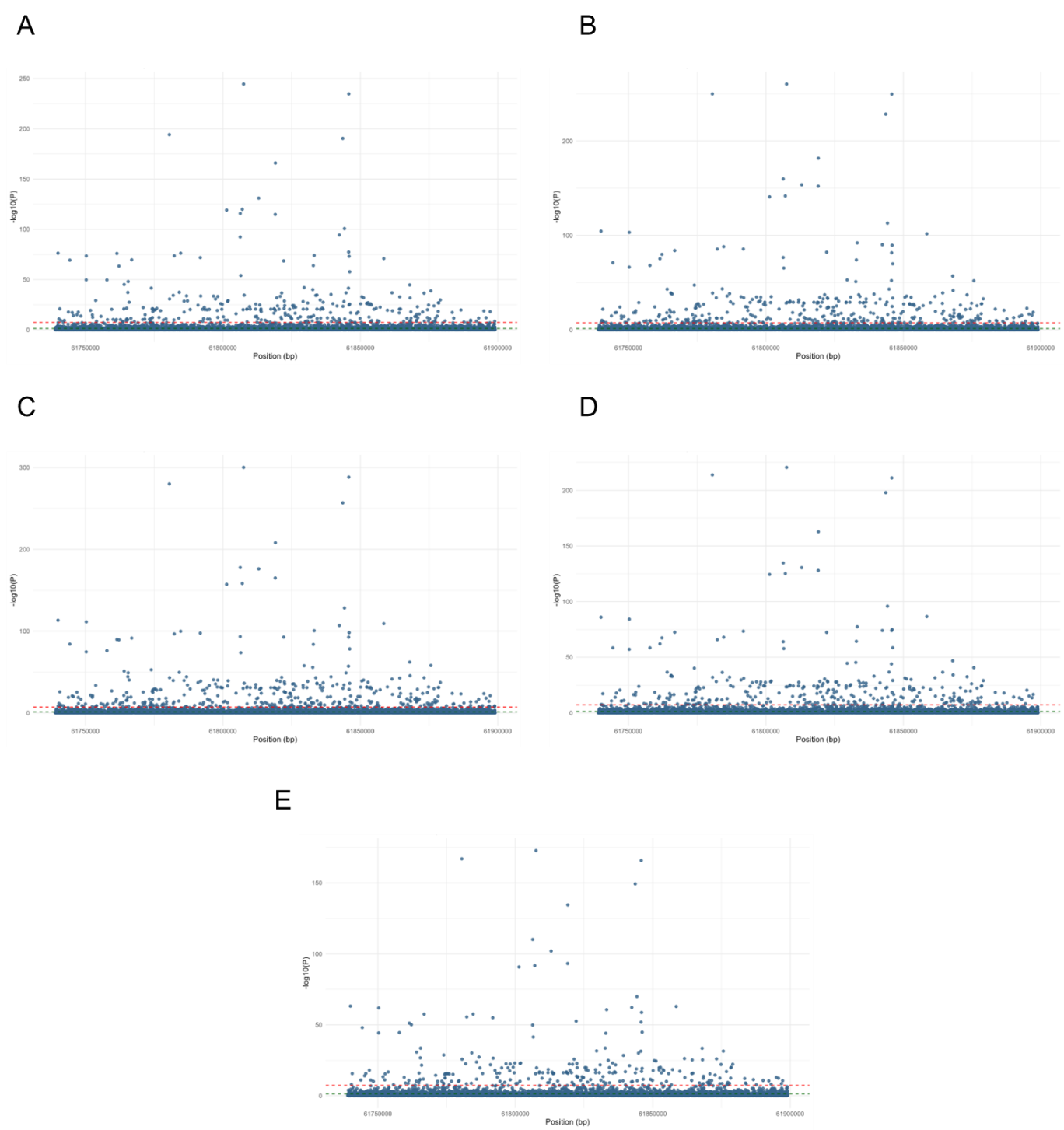

*Figure S3. Plots depicting the associations between the rare variants and metabolite levels across different masks. The black line indicates nominal significance, while the red line indicates test-wide significance. A) Unsaturation, B) Omega 6 by Omega 3, C) Omega 3 percentage, D) Omega 3, E) DHA.*

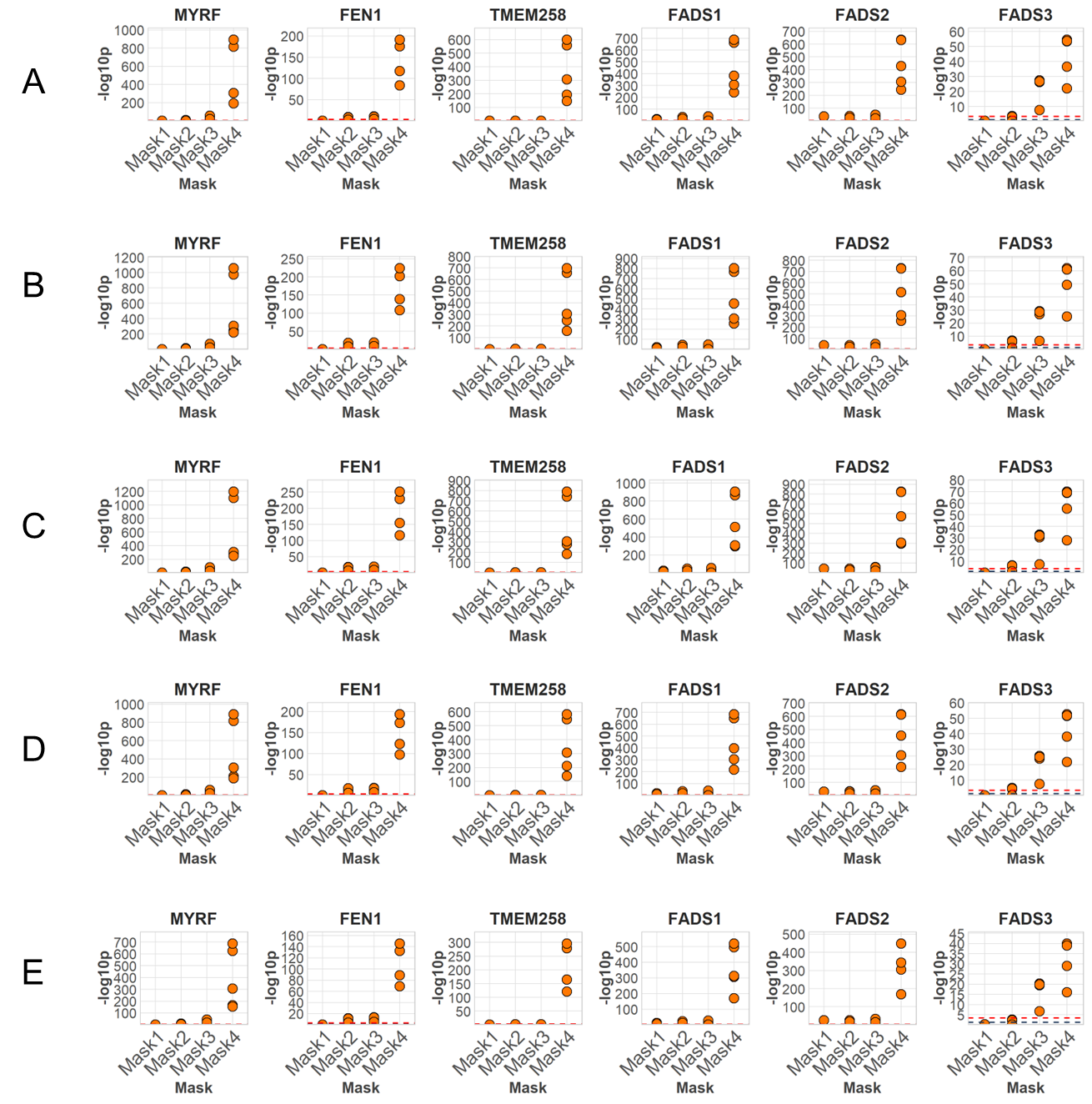

*Figure S4. Plots depicting the associations between the burden of rare variants and metabolite levels across different masks. The black line indicates nominal significance, while the red line indicates test-wide significance. A) Unsaturation, B) Omega 6 by Omega 3, C) Omega 3 percentage, D) Omega 3, E) DHA.*
